# Drug-Coated Balloon Versus Drug-Eluting Stent for Treating De Novo Large Vessel Coronary Artery Disease: A Systematic Review and Meta-Analysis of 13 Studies Involving 2,888 Patients

**DOI:** 10.1101/2023.12.10.23299794

**Authors:** Rodolfo Caminiti, Giampiero Vizzari, Alfonso Ielasi, Giampaolo Vetta, Antonio Parlavecchio, Domenico Giovanni Della Rocca, Carolina Montonati, Dario Pellegrini, Mariano Pellicano, Maurizio Tespili, Antonio Micari

## Abstract

**Introduction:** Drug-coated balloon (DCB) is an established treatment option for in-stent restenosis and small vessel, de novo, coronary artery disease (CAD). Although the use of this tool is increasing in everyday practice, little is known about its performance in the treatment of de novo, large vessel CAD (LV-CAD). A systematic review and meta-analysis were conducted to evaluate the efficacy and safety of DCB versus drug-eluting stent (DES) in this setting.

**Methods:** A comprehensive literature search was performed including Medline, Embase and Cochrane electronic databases up to October 24^th^ 2023, for studies which compared efficacy and safety of DCB versus DES in the treatment of de novo lesions in large vessels (≥ 2.5 mm), reporting at least one clinical outcome of interest (PROSPERO ID: CRD42023470417). The outcomes analysed were cardiovascular death (CVD), myocardial infarction (MI), target lesion revascularization (TLR), all cause death (ACD) and late lumen loss (LLL) at follow-up. The effect size was estimated using a random-effect model as risk ratio (RR) and mean difference (MD) and relative 95% confidence interval (CI).

**Results:** A total of 13 studies (6 randomized controlled trials and 7 observational studies) involving 2,888 patients (DCB n=1,334; DES n=1,533) with de novo LV-CAD were included in this meta-analysis following our inclusion criteria. A DCB-only strategy was non inferior to percutaneous coronary intervention (PCI) with DES in terms of CVD (RR 0.49; 95% CI [0.23 - 1.03]; p=0.06), MI (RR 0.48; 95% CI [0.16 - 1.45]; p=0.89), TLR (RR 0.73; 95% CI [0.40 - 1.34]; p=0.32), ACD (RR 0.78; 95% CI [0.57 - 1.07]; p=0.12) and LLL (MD −0.14; 95% CI [−0.30 - 0.02]; p=0.18) at follow-up.

**Conclusion:** Our meta-analysis showed that DCB PCI might provide a promising option for the management of selected, de novo LV-CAD compared to DES. However, more focused RCTs are needed to further prove the benefits of a “metal free” strategy in this subset of CAD.

## Introduction

Drug-eluting stent (DES) implantation represents the gold-standard treatment strategy for de novo CAD.^1^ However, DES are still associated with a non negligible rate of target lesion failure (TLF) at follow-up mainly due to device-related phenomena (e.g. polymer associated inflammation of the vessel wall, poor/excessive endothelialisation, incomplete stent expansion/apposition).^2,3^ In this scenario, drug-coated balloon (DCB) is emerging as a fashionable alternative to lower total stent length during PCI while preserving the anatomy and physiology of the vessel wall. A proper lesion preparation is paramount to achieve an optimal DCB PCI in order to avoid acute recoil and favour the correct penetration of the drug inside the vessel wall.^4^ Current European Guidelines recommend DCB PCI for the treatment of in-stent restenosis (ISR) with a class IA recommendation, while many clinical trials, observational studies and meta-analysis confirm its efficacy and safety in the treatment of de novo, small vessel disease (SVD).^5–7^ DCB PCI may also be considered a viable option in specific settings (e.g. high bleeding risk patients) or in association with DES in case of diffuse (e.g. long lesion/true bifurcation) CAD involving SVD.^8–10^ Although DCBs use for the treatment of de novo CAD is rapidly increasing, very little is known about the performance of a “metal free” approach for treating de novo large-vessel CAD (LV-CAD). The aim of this meta-analysis is to evaluate the efficacy and safety of DCBs compared with DES in this setting of CAD.

## Methods

### Data Sources and Searches

We systematically searched the Medline, Embase, and Scopus electronic databases for studies published until 24^th^ October 2023, focusing on those comparing the efficacy and safety of DCB and DES in the treatment of de novo LV-CAD with a reference vessel diameter (RVD) ≥ 2.5 mm and reporting at least one clinical outcome of interest. Two investigators (R.C. and G.V.) independently conducted searches using the following terms: “drug eluting stent”, “drug coated balloon”, “myocardial infarction”. Detailed information on our literature search strategy is available in the Supplementary Appendix in the Expanded Methods.

### Study Selection

The preferred reporting items for systematic reviews and meta-analyses (PRISMA) statement for reporting systematic reviews and meta-analyses was used in this study. The predefined protocol was registered to the international prospective registry of systematic reviews (POSPERO ID: CRD42023470417). Studies had to meet the following criteria in order to be included in the analysis: (1) adult (≥18 years) population; 2) direct comparison between DCB and DES, (3) ≥ 6 months clinical and/or angiographic follow-up available; (4) one or more clinical outcomes of interest reported (e.g., cardiovascular death myocardial infarction, all-cause death). Case reports, editorials, reviews, expert opinions, and studies not published in English language were excluded.

### Data Extraction and Quality Appraisal

Two investigators (R.C and G.V) extracted data from each trial using standardised protocol and reporting forms. Two reviewers (R.C and G.V) independently assessed quality items and disagreements were resolved by consensus. The Newcastle-Ottawa Quality Assessment Scale for cohort studies and the Cochrane Risk of Bias Tool for randomised clinical trials (RCTs) were used by two investigators (R.C and G.V) to assess the quality of each study.

### Study Endpoints

Cardiovascular death (CVD) was defined as death resulting from cardiovascular causes. Myocardial infarction (MI) was defined based on the fourth universal definition of PCI-Related MI ≤48 hours after the index procedure (Type 4a MI).^11^ Target lesion revascularization (TLR) was defined as a repeat PCI within the stented or DCB-treated segment or bypass surgery of the target vessel performed for restenosis or other complication of the target lesion. All cause death (ACD) was defined as death resulting from cardiovascular and other causes. The angiographic endpoint was late lumen loss (LLL) obtained by quantitative coronary angiography (QCA) and defined as minimal lumen diameter (MLD) immediately after PCI minus MLD at follow-up angiography. All endpoints were commonly defined according to the Academic Research Consortium definitions.^12^

### Statistical Analysis

Descriptive statistics are presented as mean and standard deviation (SD) for continuous variables, or number of cases (n) and percentage (%) for dichotomous and categorical variables. The Mantel-Haenszel risk ratio (RR) model was used to summarize the data for binary outcomes between treatment arms. For continuous variables, summary estimates and 95% confidence intervals (CI) were reported as the standardised mean difference. Heterogeneity between studies was assessed using the Chi2, Tau2 and Higgins-I2 statistics, and random effects models by DerSimonian and Laird were used. Subgroup analyses were performed including only RCTs.

Publication bias was assessed using funnel plots. Statistical analysis was performed with Review Manager (RevMan) (computer program) version 5.4.1, Copenhagen, Denmark: Nordic Cochrane Centre, The Cochrane Collaboration, 2020

## Results

### Study Selection and Baseline Characteristics

Among 597 screened articles, 53 full texts were retrieved and reviewed for possible inclusion; a total of 13 studies fulfilled the selection criteria and were included in the final analysis (**Figure 1**).

**Figure 1.**
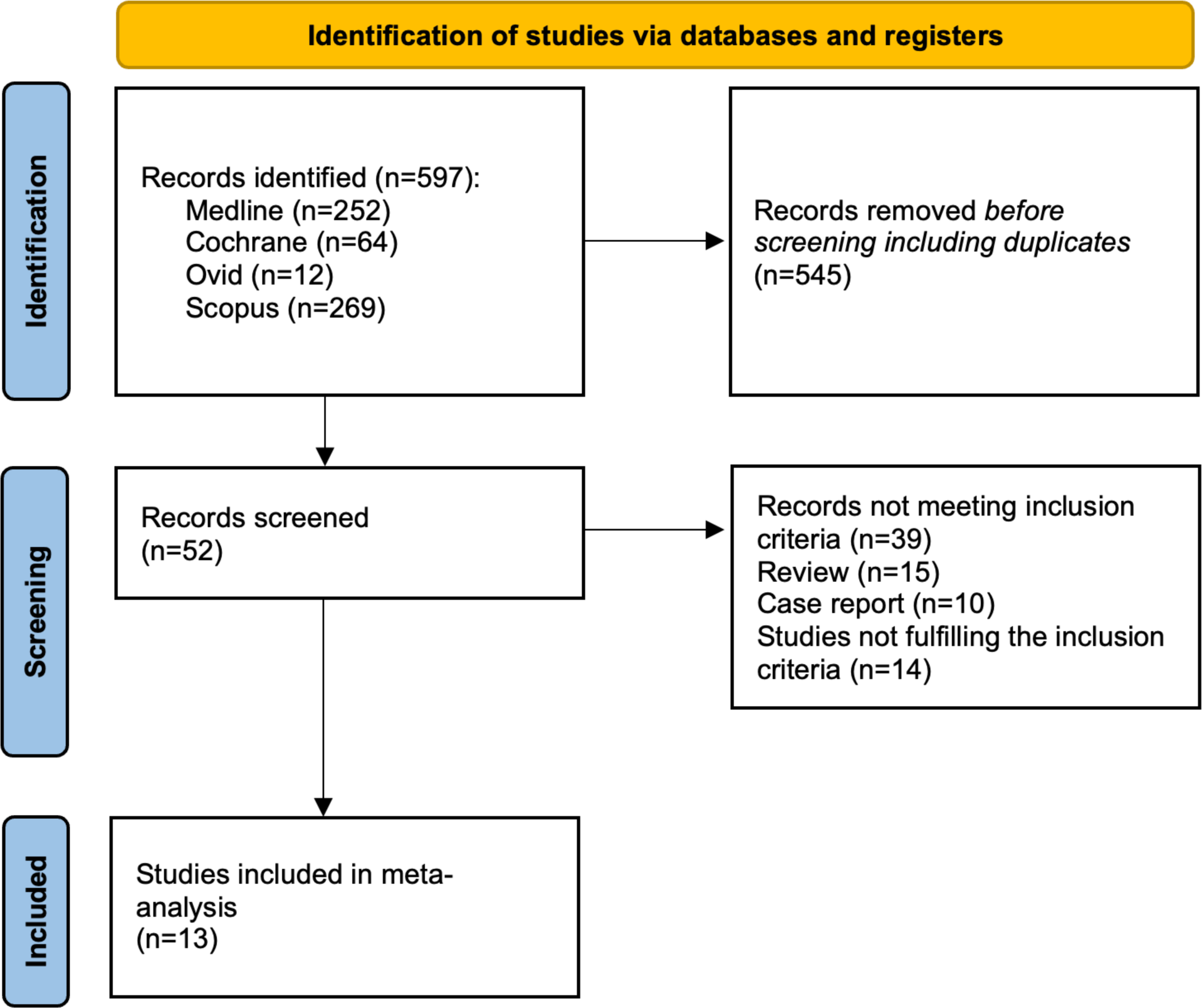
Evidence search and selection of Preferred Reporting Items for Systematic Reviews and Meta-Analyses (PRISMA)

The studies enrolled n=2,888 patients (Group DCB, n=1,334 patients; Group DES, n=1,533 patients). Overall, 75.3% (95% CI: 71.3 – 79.4%) patients were male with an average age of 63.2 years (95% CI: 57.3 - 70.5). The indication for revascularization was in 60.2% (95% CI: 38.7 – 85.1%) of cases acute coronary syndrome (ACS). The left anterior descending (LAD) artery was treated in the majority of cases (47.1%; 95% CI: 35.8 – 57.9%), followed by the right coronary artery (25.5%; 95% CI: 19.9 – 33.7%), left circumflex artery (18.1%; 95% CI: 11.5 – 23.3%) and unprotected left main (ULM) (9.3%; 95% CI: 6.1 – 23.2%).

The mean lesion length was 22.8 mm (95% CI: 15.3 - 40.2 mm) in the DCB and 27.9 mm (95% CI: 18.1 - 45.6 mm) in the DES group. The mean reference vessel diameter (RVD) was 3.14 mm (95% CI: 2.79 – 3.32 mm) in the DCB and 3.18 mm (95% CI: 2.69 – 3.57 mm) in the DES group.

All studies used paclitaxel coated balloons (PCB) except one in which sirolimus coated balloon (SCB) was also used.

Further details on baseline characteristics, clinical and angiographic follow-up time of the studies population are reported in **Table 1**.

**Table 1.**
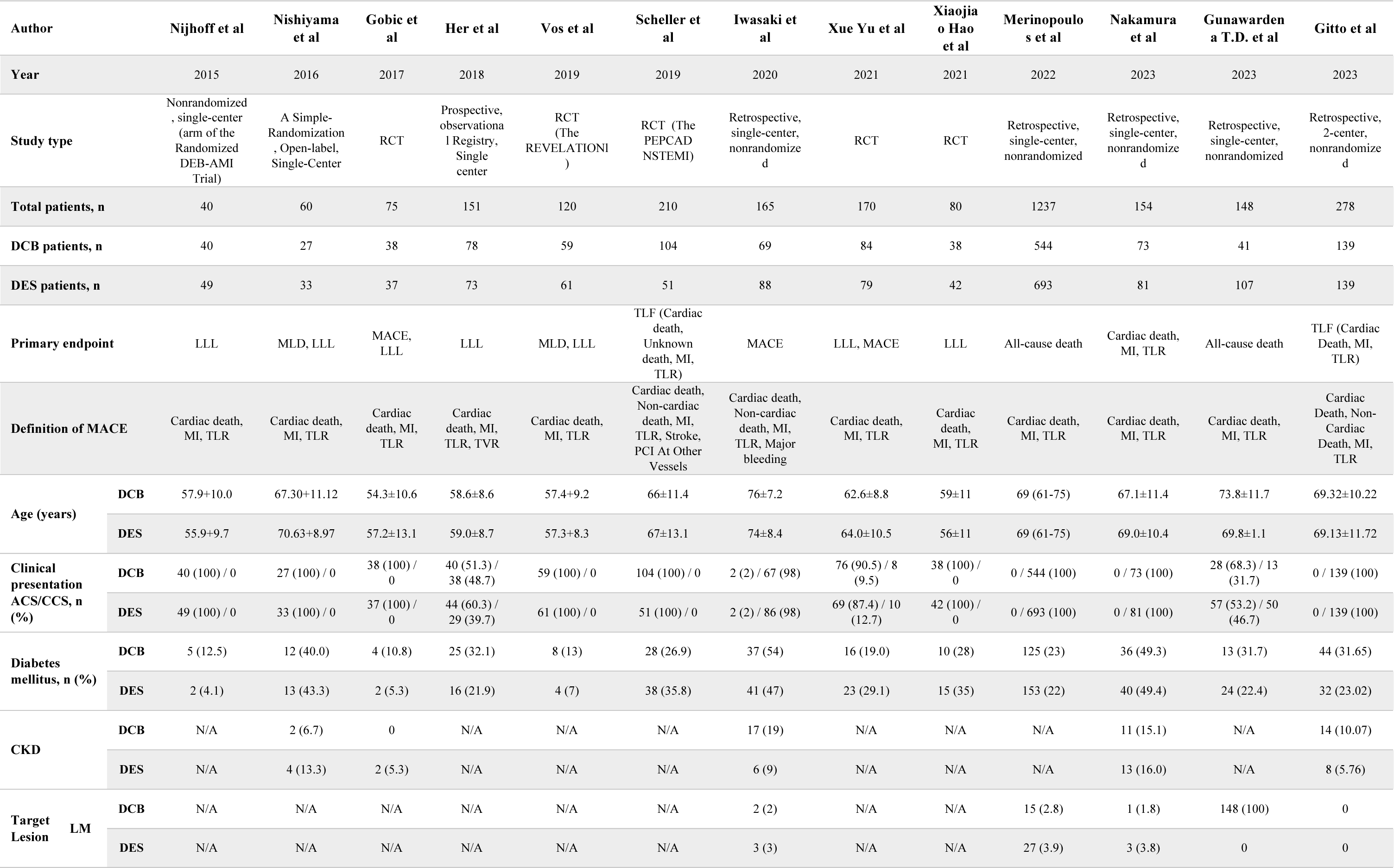
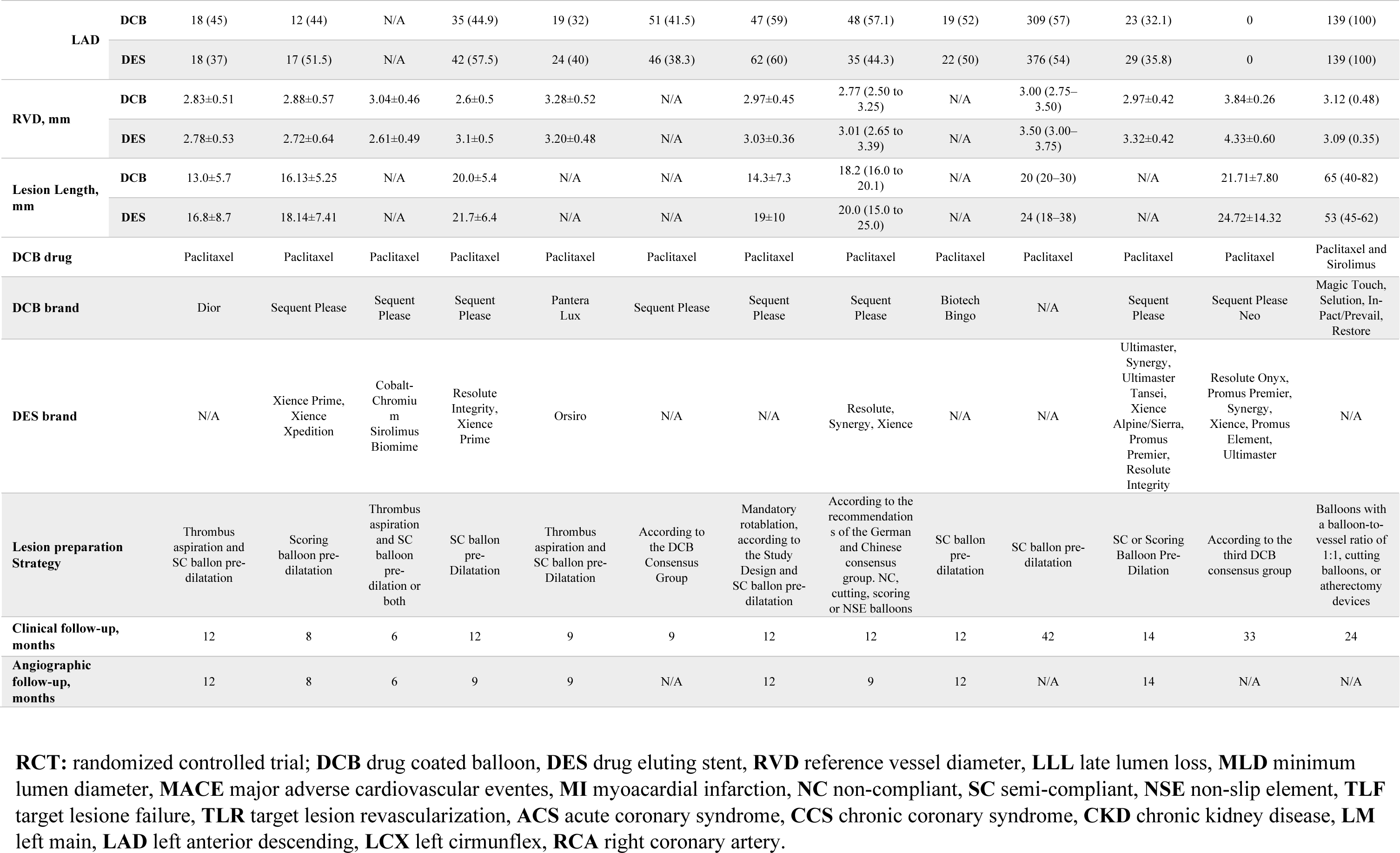
Main Features of the Studies Included in the Meta-Analysis.

### Endpoints

Twelve studies reported clinical follow-up data on CVD, MI and TLR.^13–24^ No differences were found between DCB and DES for the risk of CVD [1.1% vs 3.2%; RR: 0.49 (95% CI: 0.23 - 1.03; p=0.06; I^2^=0%] (**Figure 2**); MI [0.3% vs 1.5%; RR: 0.48 (95% CI: 0.16 - 1.45; p=0.89; I^2^=0%] (**Figure 3**), and TLR [3.7% vs 6.1%; RR: 0.73 (95% CI: 0.40 - 1.34; p=0.32; I^2^=27%] (**Figure 4**). Eight studies reported data on ACD^14,16–20,24,25^. At follow-up, no differences were found between DCB and DES for the risk of ACD [5.5% vs 7.8%; RR: 0.78 (95% CI: 0.57 - 1.07; p=0.12; I^2^=0%] (**Figure 5**).

**Figure 2.**
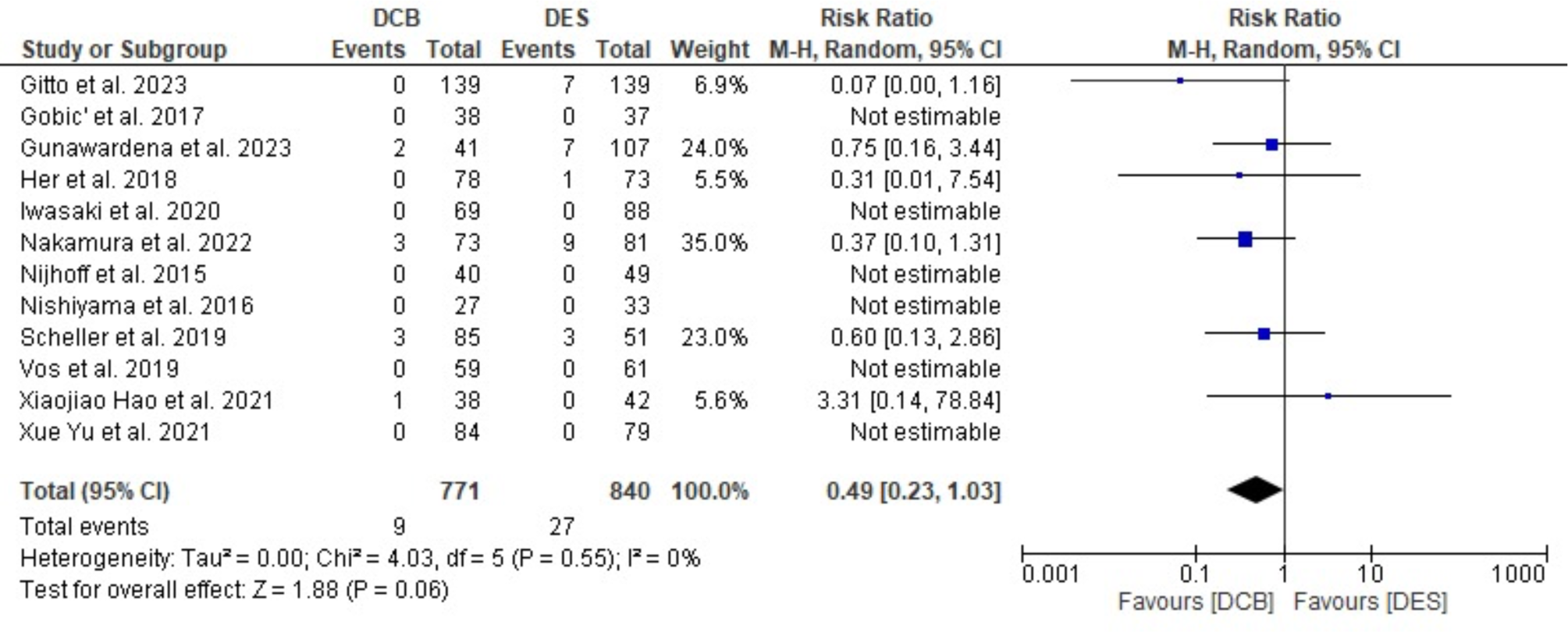
Forest plots comparing Cardiovascular Death between Drug Coated Balloon Versus Drug Eluting Stent.

**Figure 3.**
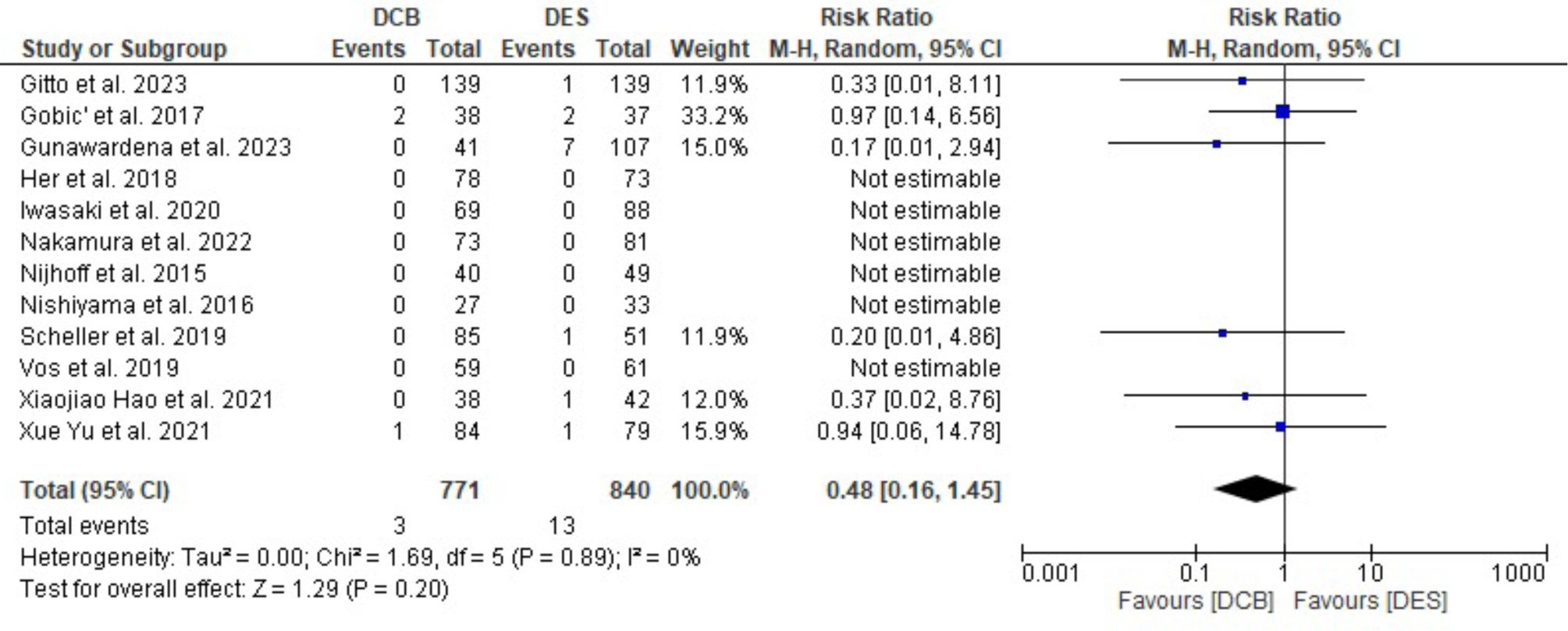
Forest plots comparing Myocardial Infarction between Drug Coated Balloon Versus Drug Eluting Stent.

**Figure 4.**
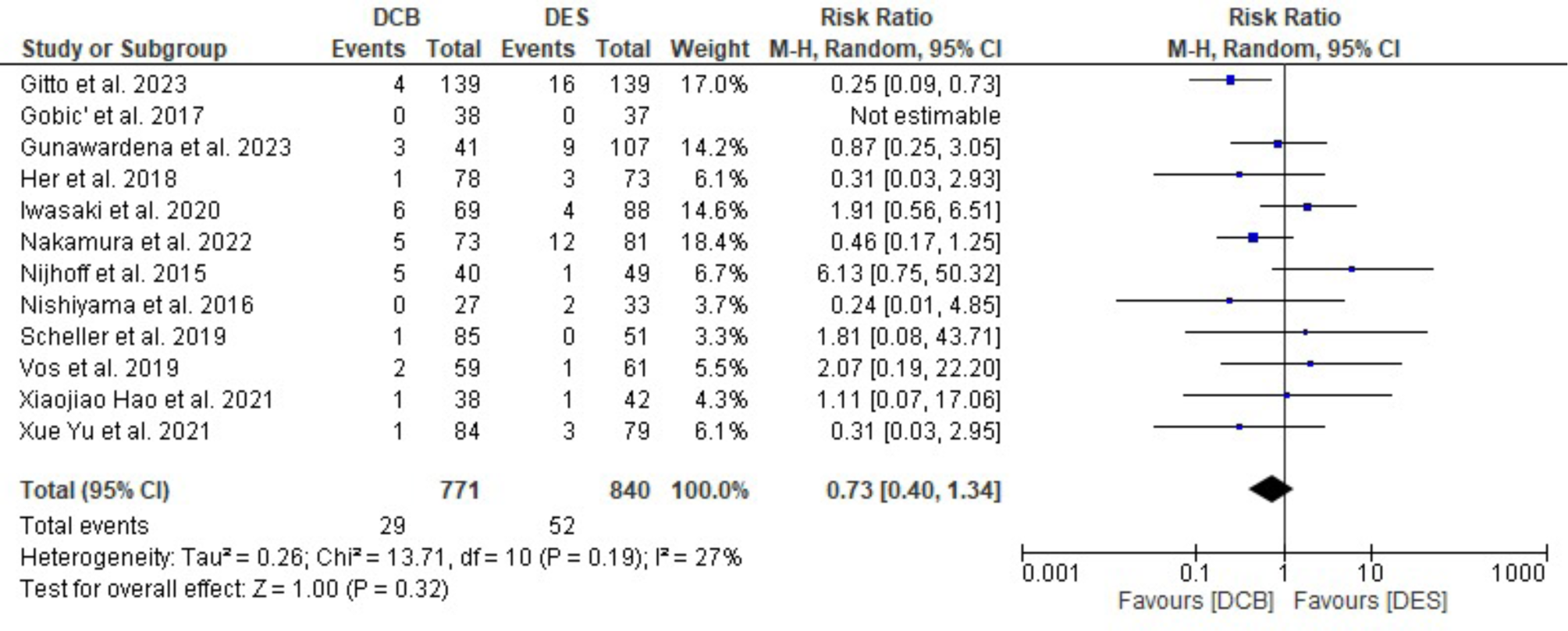
Forest plots comparing Target Lesion Revascularization between Drug Coated Balloon Versus Drug Eluting Stent.

**Figure 5.**
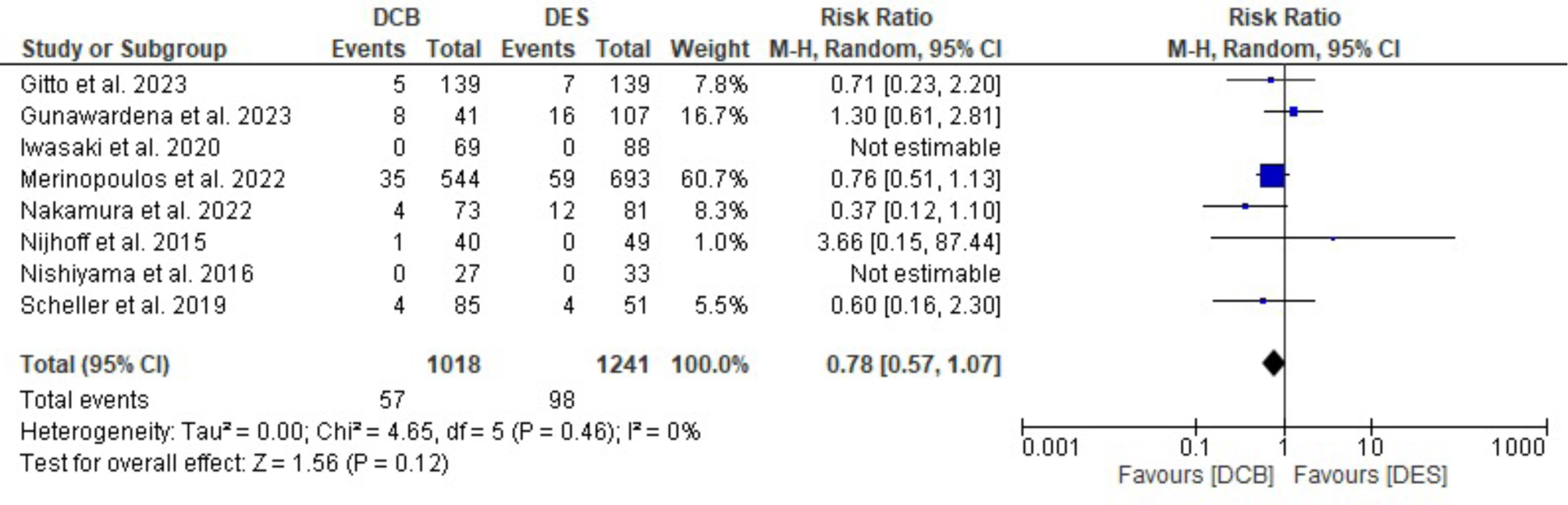
Forest plots comparing All Cause Death between Drug Coated Balloon Versus Drug Eluting Stent.

In terms of angiographic results, nine studies reported data on LLL.^13,15–19,21–23^ DCB was not inferior to DES for LLL at follow-up [MD: −0.14 (95% CI: −0.30 - 0.02; p=0.10; I^2^=91%] (**Figure 6**). Finally, six studies reported data on MLD before and after PCI.^15–19,23^ DES proved a higher mean acute gain versus DCB [1.94 (1.73, 2.14) vs 1.31 (1.02, 1.60); p=0.0006; I^2^ 91.6%] (**Figure 7**).

**Figure 6.**
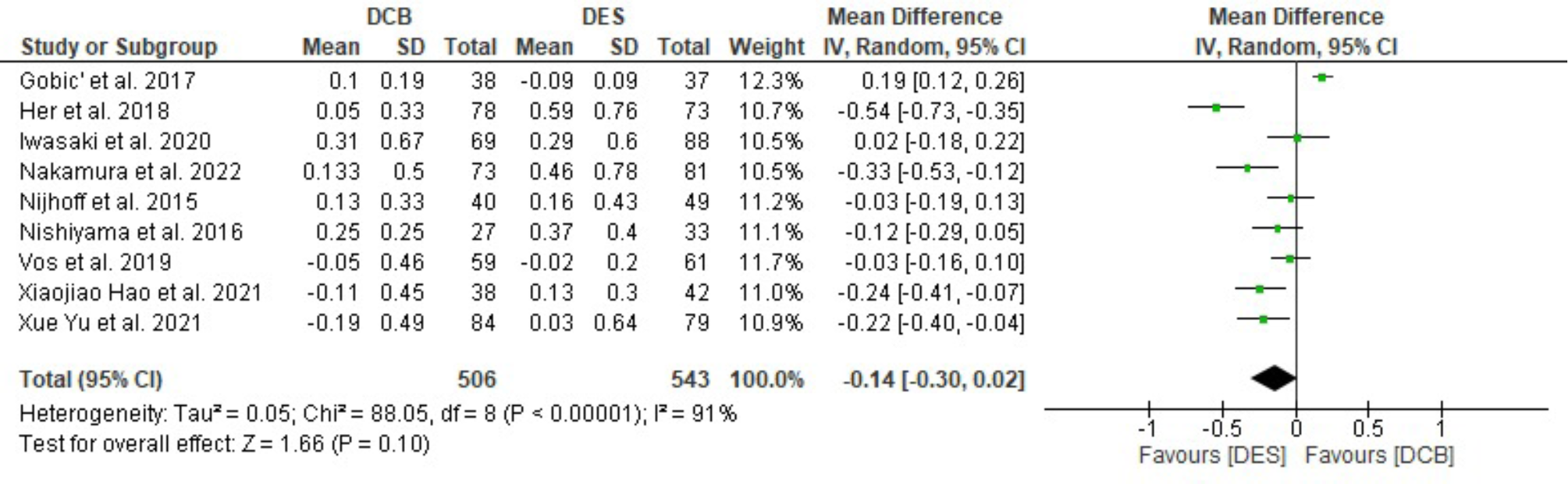
Forest plots comparing Late Lumen Loss between Drug Coated Balloon Versus Drug Eluting Stent.

**Figure 7.**
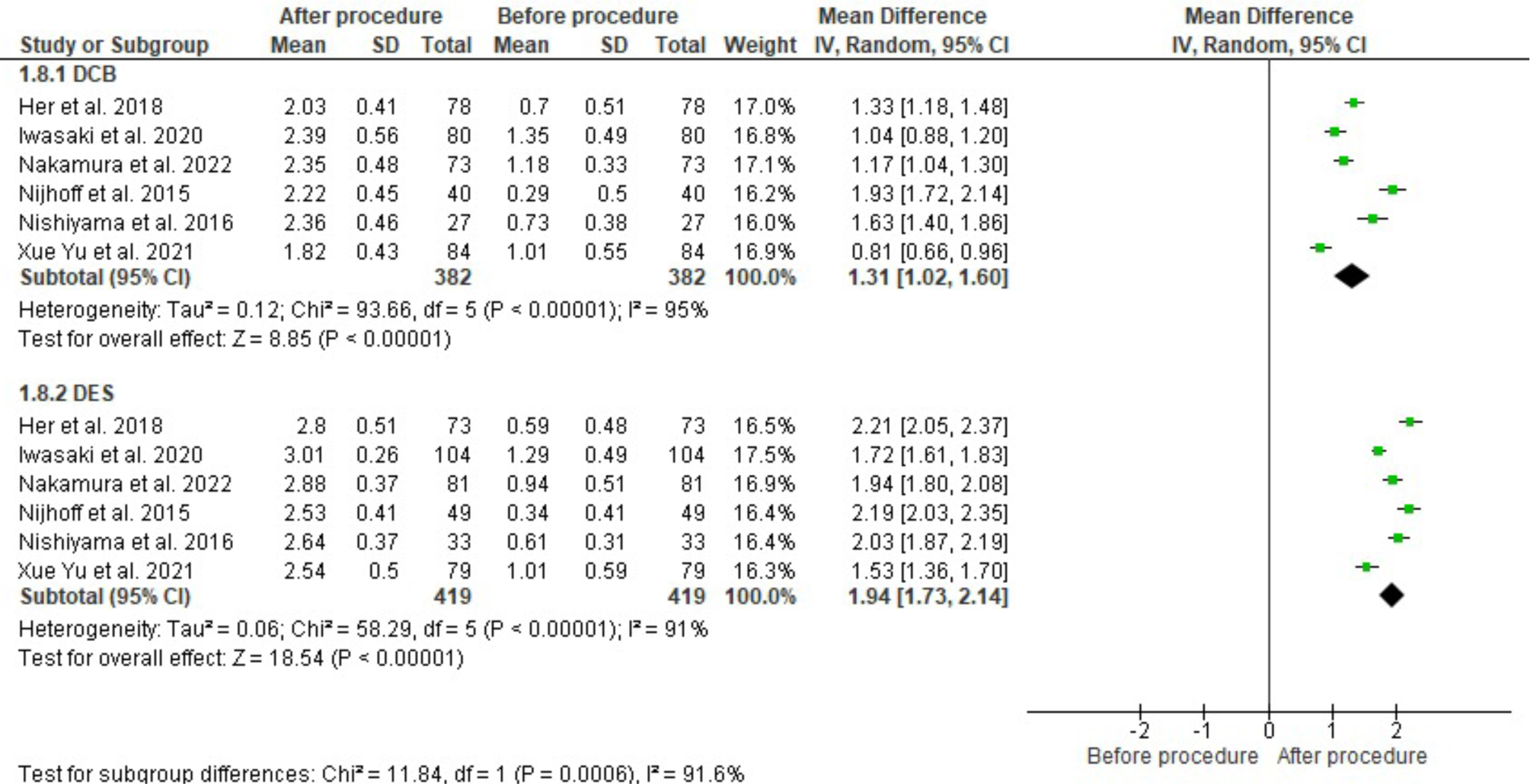
Forest plots comparing Minimal lumen diameter before and after procedure between Drug Coated Balloon Versus Drug Eluting Stent.

### Subgroup analysis including only RCTs

Six RCTs reported data on CVD, MI and TLR.^13,19–23^ At follow-up, no differences were found between DCB and DES for the risk of CVD [1.2% vs 0.9%; RR: 0.84 (95% CI: 0.21-3.40; p=0.80; I^2^=0%] (**Figure 8A**), MI [0.9% vs 1.6%; RR: 0.64 (95% CI: 0.18-2.30; p=0.49; I^2^=0%] (**Figure 8B**) and TLR [1.5% vs 2.3%; RR: 0.77 (95% CI: 0.24-2.50; p=0.67; I^2^=0%] were found (**Figure 8C**). Two RCTs reported data on ACD.^19,20^ No differences were found between DCB and DES for the risk of ACD at follow-up [3.5% vs 4.7%; RR: 0.60 (95% CI: 0.16-2.30; p=0.46; I^2^=0%] (**Figure 8D**).

**Figure 8.**
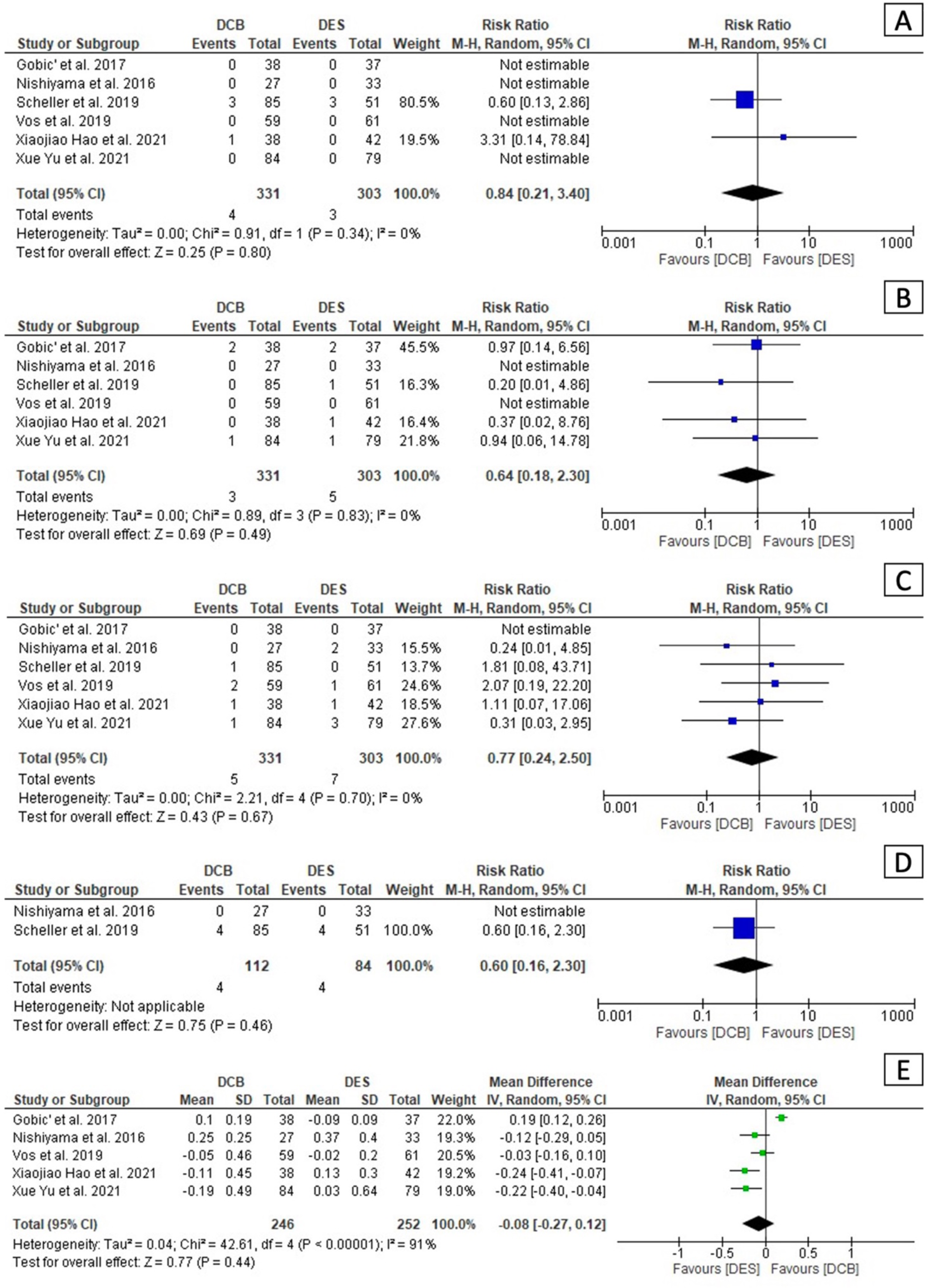
Forest plots comparing Cardiovascular Death (A), Myocardial Infarction (B), Target Lesion Revascularization (C), All Cause Death (D) and Late Lumen Loss (E) between Drug Coated Balloon Versus Drug Eluting Stent in randomized controlled trials subgruoup.

Six RCTs studies reported data on LLL.^13,19,21–23^ No differences were observed between DCB and DES for LLL at follow up [MD: −0.08 (95% CI: −0.27 - 0.12; p=0.44; I^2^=91%] (**Figure 8E**).

Two RCTs studies reported data on MLD before and after the procedure.^19,23^ DES proved a higher MLD mean difference before and after PCI [1.79 (1.67, 1.91) vs 1.06 (0.94, 1.18); p<0.00001; I^2^=98.6%] (**Figure 9**).

**Figure 9.**
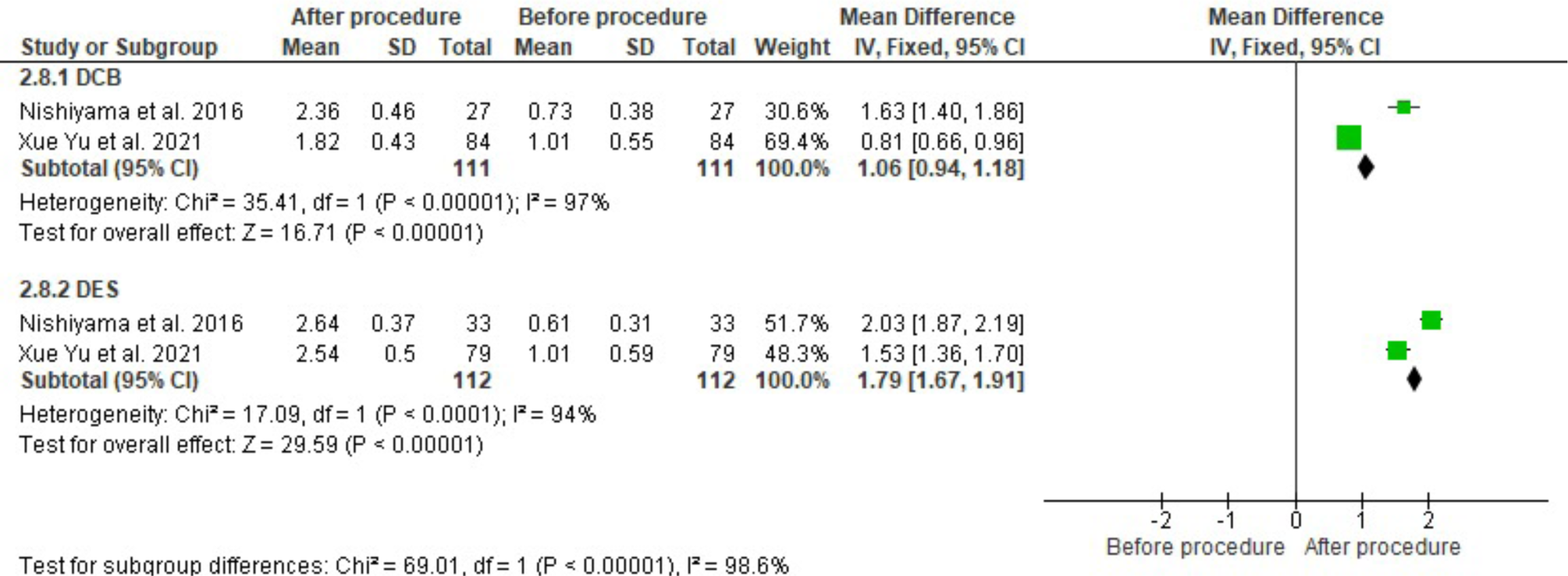
Forest plots comparing Minimal Lumen Diameter before and after the procedure between Drug Coated Balloon Versus Drug Eluting Stent in randomized controlled trials subgruoup.

### Publication Bias

A graph and summary of Cochrane Risk of Bias tool for RCTs and Newcastle–Ottawa Quality Assessment Scale for cohort studies is reported in **Supplemental Figure 1**. The funnel plots for visual inspection of the bias showed no bias (**Supplemental Figure 2**).

## Discussion

In this meta-analysis, we evaluated the role of a “metal-free” approach with DCB for the treatment of de novo LV-CAD in both, acute and elective settings.

In summary, our results suggest that the usage of DCB-only strategy in this scenario is safe and effective with similar clinical and angiographic results compared to DES. The feasibility of treating LV-CAD with a DCB was initially established in retrospective and prospective observational registries.^26–28^

Several RCTs assessed the role of a “metal free” approach vs. DES PCI in selected de novo lesions (e.g. “culprit”, heavily calcified and ULM) located in large vessels. Consistently with other studies, our analysis confirmed that, in LV-CAD PCI, DES is associated with a significantly higher acute gain as compared to DCB (1.94 mm vs 1.31 mm; p=0.0006). However, LLL at follow-up was similar between the devices, claiming for a positive remodelling related to DCB PCI.^4,5,29,30^

In a sub-analysis of the DEB-AMI trial, Nijhoff et al demonstrated the feasibility of a DCB-only PCI (n=40) in ACS with large target vessel disease (average RVD: 2.83 ± 0.51 mm, lesion length: 13.0 ± 5.7 mm, DCB diameter: 3.3 ± 0.4 mm). Despite the higher LLL at 12-month follow-up in the DCB arm (0.51 ± 0.59 mm vs. 0.21 ± 0.32 mm, p<0.01), no differences in ACD, target-vessel revascularization (TVR), TLR, MI were reported as compared to DES.^18^

The first randomized head-to-head comparison of DCB vs. DES in STEMI patient with LV-CAD (average RVD 2.61 ± 0.49 mm DCB group, 3.04 ± 0.46 mm DES group) reported comparable LLL and major adverse cardiovascular events (MACE: a composite of CVD, re-MI, TLR, and stent thrombosis) rate at 6-month follow-up between the groups (LLL 0.10 ± 0.19 mm DES vs. −0.09 ± 0.09 DCB, p<0.05).^13^

The REVELATION trial evaluated the performance of a DCB-only PCI in STEMI patients (n=120) undergoing primary PCI on a large culprit vessel (average RVD 3.28 ± 0.52 mm DCB group, 3.20 ± 0.48 mm DES group). Primary end-point was the fractional flow reverse (FFR) value at the end of the procedure and at 9-month follow-up. No major adverse events occurred at follow-up and the angiographic/functional outcomes were similar between the two groups (mean FFR 0.92 ± 0.05 DCB vs. 0.91 ± 0.06 DES, p=0.27; mean LLL −0.05 ± 0.46 mm DCB vs. −0.02 ± 0.2 mm DES, p=0.51).^21^

The PEPCAD NSTEMI trial randomized 210 patients comparing DCB vs. stent [bare metal stent (BMS) and DES] in de novo, non-thrombus containing LV-culprit lesions (RVD as inclusion criteria: 2.5-3.5 mm). No differences were found between the groups in terms of CVD/ACD, re-MI, and TLR at 9-month follow-up (DCB: 4.7% vs. DES: 5.9%, p<0.88).^20^

An interesting point arising from these studies relates to the usage of DCB PCI in “culprit” lesions, which are not considered a good spot for a metal-free approach because of the potentially lower dose of drug transferred to the vessel wall, particularly in thrombus containing lesions. Indeed, the sub-analysis of the DEB-AMI trial showed a higher LLL in the DCB arm. However, this result might have been influenced by the DCB used (the first generation DIOR delivers 25% only of the drug dose to the vessel wall).^31^

DCB PCI was also challenged vs. DES in heavily calcified lesions requiring rotational atherectomy (average RVD 2.97 ± 0.45 mm, lesion length 14.3 ± 7.3 mm DCB group; average RVD 3.03 ± 0.36 mm, lesion length 19 ± 10 mm DES group). Angiographic and clinical outcomes at 1-year follow-up (LLL 0.29 ± 0.60 mm DES vs. 0.31 ± 0.67 mm DCB, p=0.83; MACE 8% DES vs. 11% DCB, p=0.30) were similar between the groups.^16^

Even in de novo ULM disease (average RVD 3.84 ± 0.26 mm, lesion length: 21.71±7.80 mm DCB group; average RVD 4.33 ± 0.60 mm, lesion length 24.72 ± 14.32 mm DES group) DCB PCI was associated with similar results as compare to DES [TLR: 7.3% DCB vs. 8.3% DES (HR: 0.89 (0.24–3.30), p=0.86)] at a median of 33 months follow-up.^14^

More recently, a propensity score (PS) matching analysis of a DCB-alone or in combination with DES (“hybrid” strategy) versus a DES-alone strategy (n=139 pairs) in the treatment of de novo long LAD lesions (mean RVD 3.14 ± 0.40 mm, lesion length 56.87 ± 16.7 mm) resulted in a lower TLF rate (TLR, CVD, and target vessel-MI) at 2 years in the DCB group as compared to the DES group (5.9% DES vs. 0% DCB). Furthermore, a signal toward lower CVD risk was reported in the DCB group. This finding is consistent with the results of our meta-analysis, where this outcome is close to significance (p=0.06), hypothesizing an advantage of the “metal-free” approach.^24^

Looking at technical aspects, pre-dilation was mandatory in all treatment groups. This manoeuvre is a key step for a successful PCI, particularly in case of DCB usage. According to the DCB consensus group, an optimal lesion preparation (e.g. residual % diameter stenosis less than 30) is required prior to DCB inflation.^32^ An “aggressive” (e.g., non-compliant -NC-balloons escalation to super high-pressure NC balloons, scoring/cutting balloons, intravascular lithotripsy, debulking devices) pre-dilatation strategy could facilitate plaque incision and drug transfer to the vessel wall, reducing elastic recoil and influencing a good clinical outcome.^4^

Even in ACS setting and in presence of thrombus, optimal lesion preparation is mandatory before inflating a DCB. Proper thrombus aspiration, which was performed in many of the ACS patients enrolled in the above studies (78% in the DCB group of the REVELATION Trial), could be crucial to reduce the number of pre-dilatation balloon inflations and the subsequent risk of distal embolization while facilitating drug penetration in the vessel wall.

Although vessel preparation plays a key role in DCB PCI, on the other hand may be associated with vessel injuries. Indeed, a main concern associated with DCB-PCI-only in proximal LV-CAD is the occurrence of malignant dissections. Cortese et al. assessed the fate of leaving non-flow-limiting dissection (A-C) after DCB PCI. At 6-month angiographic follow-up complete vessel healing was reported in 93.8% of cases while a low incidence of major adverse events occurred at 9-month follow-up (7.2% no-dissection cohort vs. 8.1% dissection cohort; p=0.48). The authors hypothesised that paclitaxel may play a role in facilitating coronary vessel healing when properly delivered at the target site.^33^ Beside to angiography, a functional evaluation could lead the management of a dissection in the setting of DCB PCI. Especially in case of type A-B dissection, a Pd/Pa threshold of more or equal than 0.90 may be used as a surrogate for optimal outcome (leaving the dissection) while a Pd/Pa less than 0.90 may lead to bail-out DES implantation reducing the risk of abrupt vessel closure and MI.^34^

Of note, most of the studies included in this meta-analysis assessed the performance of PCBs, with the most commonly used brand being Sequent Please (B. Braun) in seven studies.

The main difference among PCBs is related to the formulation of the water-soluble excipient and the drug concentration, with the first aspect mostly influencing the final effect on the vessel wall, due to its sustained release properties. Although a non-randomised, score-matched comparison (SIRPAC trial) of two large registries assessing the performance of a PCB (Elutax SV, Aachen Resonance, Lainate, Italy) versus a first generation SCB (Magic Touch, Concept Medical, Tampa, Florida, USA) reported similar clinical results at 12 months^35–37^, a recent randomized study showed that the same SCB resulted inferior to SeQuent Please Neo PCB in terms of angiographic net lumen gain at 6 months (lumen gain 0.25 ± 0.40 mm SCB, 0.48 ± 0.37 mm PCB).^38^ These results deserve further attention particularly when choosing a DCB in the setting of LV-CAD.

Last but not least, the studies included in this meta-analysis are mainly related to selected lesions (length in both arms less than 28 mm) in specific subsets (e.g. calcified, ULM and ACSs), which are currently considered “off-label” for DCBs usage. Despite that, our results showed encouraging outcomes associated with the “metal-free” approach. Data from ongoing RCTs comparing current generation DCBs vs. DES in large cohort of patients including those with LV-CAD are awaited.^39,40^

## Limitations

Main limitation of our meta-analysis is the inclusion of observational studies beside to RCTs, which might limit the strengthens of the study. However, our results were confirmed at the subgroup analysis of RCTs only.

## Conclusions

DCBs are an attractive option for the treatment of de novo CAD. Our meta-analysis showed no significant clinical and angiographic differences between DCB and DES in treating LV-CAD in either acute or elective settings. Focused RCTs providing further evidence on the potential benefit of a metal free approach in LV-CAD are strongly needed.

## Data Availability

The data underlying this article are available in the article and in its online supplementary material.

## Acknowledgments

none

## Abbreviations

ACD: All cause death
CAD: Coronary artery disease
CI: Confidence interval
CVD: Cardiovascular death
DAPT: Dual antiplatelet therapy
DCB: Drug-coated balloon
DES: Drug-eluting stent
ISR: In-stent restenosis
LV: Large vessel
MACE: Major adverse cardiovascular events
MD: Mean difference
MI: Myocardial infarction
MLD: Minimal lumen diameter
PCB: Paclitaxel coated balloon
PCI: Percutaneous coronary intervention
PRISMA: Preferred reporting items for systematic reviews and meta-analyses
QCA: Quantitative coronary analysis
RR: Risk ratio
SCB: Sirolimus coated balloon
SVD: Small vessel disease
TLF: Target lesion failure
TLR: Target lesion revascularization
ULM: Unprotected left main

